# An optimized flow cytometry protocol for simultaneous detection of T cell activation induced markers and intracellular cytokines: application to SARS-Cov-2 vaccinated individuals

**DOI:** 10.1101/2022.04.14.22273819

**Authors:** Tiziana Altosole, Gianluca Rotta, Scott J. Bornheimer, Daniela Fenoglio

## Abstract

Antigen (ag)-specific T cell analysis is an important step for investigation of cellular immunity in many settings, such as infectious diseases, cancer and vaccines. Multiparameter flow cytometry has advantages in studying both the rarity and heterogeneity of these cells. In the cellular immunologist’s toolbox, the expression of activation-induced markers (AIM) following antigen exposure has made possible the study and sorting of ag-specific T cells without using human leukocyte antigen (HLA)-multimers. In parallel, assessing the cytokine profile of responding T cells would support a more comprehensive description of the ongoing immune response. Here, a method and flow cytometry panel were optimized to combine the detection of activated CD4+ and CD8+ T cells in a TCR-dependent manner with the evaluation of cytokine production by intracellular staining, without affecting the positivity of activation markers. In particular, the expression of CD134 (OX40) and CD69 have been tested in conjunction with intracellular (ic) CD137 (4-1BB) to detect SARS-Cov-2 Spike protein-specific activated T cells. In our setting, assessing CD134 provided minimal contribution to detect the pool of AIM+ T cells, whereas a key role was described for ic-CD69, which was co-expressed with ic-CD137 in both CD4+ and CD8+ lymphocytes. Moreover, the analysis of TCR-triggered cytokine-producing T cells (IFNγ, TNFα and IL-2 were assessed) further confirmed the capacity of ic-CD69 to identify functionally responsive antigen-specific T cells, which were often largely negative or poorly positive for CD134 expression. In parallel, the use of CD45RA, CCR7 and CXCR5 allowed us to describe the T cell matuarion curve and detect T helper follicular CD4+ cells, including the antigen-specific activated subsets.

In conclusion, we optimized a method and flow cytometry panel combining assessment of activation induced markers and intracellular cytokines that will be useful for measuring TCR stimulation-dependent activation of CD4+ and CD8+ T cells.

## INTRODUCTION

The identification and study of ag-specific T cells is key for a broad range of immunological research, including infectious diseases and vaccine responses (Appay et al., 2008). T cell mediated immunity against a pathogen or following vaccine treatment is a consequence of the coordinated contribution of two arms: cytotoxic CD8+ T cells mainly responsible for the clearance of target cells (Van Lier et al., 2003) and CD4+ T cells that interact with B cells and CD8+ T cells for optimal development, maturation, and maintenance of immunity against pathogens or vaccines (Borst et al., 2018). Therefore, monitoring both CD4+ and CD8+ responses is crucial for immunological research. Peptide/HLA (pHLA) multimers (Altman et al., 1996) accurately detect ag-specific T cells, but the knowledge of the immunogenic peptide, its HLA allele restriction, and the availability of the reagents are limiting factors, in particular for the CD4+ T cell population, and rarely sufficient to appreciate the breadth of ag-specific T cells in relation to functional capabilities. Deep analysis of the ag-specific T cell repertoire requires assays that focus on specific functions such as production of a particular cytokine or degranulation in response to antigen (Manz et al., 1995), (Betts et al., 2003). However, such functional assays often reflect only a subset of specifically activated cells, as the in vivo T-cell response is composed of functionally diverse cells that cannot be identified using a single cytokine or the detection of degranulation (Mosmann et al., 1997), (Sallusto et al., 2004). To overcome the limitations of these approaches and discriminate simultaneously both TCR-stimulated CD4+ and CD8+ T cells, the detection of activation-induced markers (AIMs) in response to antigen would facilitate the assessment of the complete antigen-specific T cell response.

Favorable characteristics of such surrogate markers would include specific surface expression after activation over a transient but sufficiently prolonged time period to allow reliable detection, corroborated by absent or low molecule expression if unstimulated and during resting phases. Several markers have been shown to be specifically induced in a TCR stimulation-dependent manner, such as CD154, CD107, CD137 and CD134 (Betts et al., 2003), (Chattopadhyay et al., 2005), (Chattopadhyay et al., 2006), (Frentsch et al., 2005), (Reiss et al., 2017), (Wehler et al., 2008), (Zaunders et al., 2009). CD154 (CD40L) molecule is mainly expressed on activated CD4+ T cells, being in fact proposed to monitor and isolate antigen-specific T helper cells (Chattopadhyay et al., 2006), (Meier et al., 2008), (Yellin et al., 1994). CD107A is an endosomal component and translocates to the cell surface during degranulation of antigen-specific T cells triggered by TCR stimulation; therefore, it is considered the most significant marker to define the cytotoxic potential (Betts et al., 2003), (Rubio et al., 2003). CD137 (4-1BB) is a member of the TNF Receptor family that mediates costimulatory function, and it has been identified as an up-regulated molecule expressed on activated human CD8+ and CD4+ T cells from 12 hours to up to 5 days after stimulation depending on the stimulus (Vinay and Kwon, 1998), (Watts, 2005), (Cannons et al., 2001), (Lee et al., 2002), (Dawicki and Watts, 2004). CD134 (OX-40) also belongs to the TNF Receptor superfamily and is a marker of antigen-specific CD4+ T cells (Zaunders et al., 2009). More recently, the CD134 and CD137 combined expression has been used to identify antigen-specific CD4+ T cells (Reiss et al., 2017), (Dan, 2017).

Moreover, early activation markers such as CD25 and CD69 have been used to identify antigen responsive CD4+ T cells. Even if these markers assessed alone may lack specificity (Bremser et al., 2015), (Kmieciak et al., 2009), their adoption in conjunction with TCR stimulation dependent markers may increase the sensitivity and discrimination of rare cells in flow cytometry data.

Among the TCR stimulation dependent markers, CD137 appears to be particularly interesting given its kinetics of expression after antigen recognition and its biologic role to provide a survival signal to activated T cells, both CD4+ and CD8+ T lymphocytes (Watts, 2005), (Wen et al., 2002). In parallel, emerging evidence points to the value of assessing the cytokine profile of AIM+ T cells to better describe the intensity and features of ongoing immune response (Painter MM., 2021), (Braun et al., 2020), (Yu et al., 2021).

In the present study, we optimized a straightforward protocol and related flow cytometry panel for detection of ag-specific CD4+ and CD8+ T cells. This was developed in the context of SARS-Cov-2 Spike protein specificity in individuals having received the BNT162b2 mRNA vaccine. The study resulted in a method relying on intracellular evaluation of activation-induced CD137 and CD69 expression in combination with Th1 cytokine profiling that allows describing the antigen-specific T cell maturation curve in addition to the T helper follicular cell sub-population.

## RESULTS

To recover the maximum number of TCR-dependent activated T cells and in parallel evaluate their cytokine profile, three points have been addressed in optimizing the protocol: the impact of cytokine release inhibitors on AIM expression, the combination of AIM markers, and the flow cytometry gating and data analysis approach.

A key consideration to optimize a reliable AIM test combined with cytokine detection is whether AIM marker staining should be performed on the cell surface (s) or intracellular (ic), since inhibitors of endosomal trafficking (Brefeldin A BFA and monensin) are known to prevent secretion of both cytokines and surface markers (Nylander et al., 1999), (O’Neil-Andersen et al., 2002). Our previous experience (not shown), in agreement with published literature, suggests that ic-CD137 staining can be performed successfully, in parallel with ic-cytokine evaluation, to detect both CD4+ and CD8+ T cell activation. This can be done after overnight incubation in the presence of stimuli and inhibitor of endosomal trafficking (BFA) (Yan et al., 2017), (Braun et al., 2020).

### Gating strategy

In addition, the appropriate gating strategy is crucial for succesful detection of TCR-dependent activated T cells. During the analysis phase of the dataset generated in these conditions, it emerged that activated ic-CD137bright T cells underwent a 60% statistically significant decrease of CD3 Mean Fluorescent Intensity (MFI) (p=7.6^-19^). This caused loss of 30% of events in stimulated samples when T cells were gated by the classical CD3 vs SSC dot plot. Notably, CD8 and CD4 staining were also affected by a statistically significant 32% and 6% of MFI decrease respectively (p=8.8^-15^ and p=0.003), however these changes were not sufficient to position these cells out of the classical CD4+ or CD8+ gates. (Supplemental figure 1A, B, C). As such, we decided to optimize the gating strategy for CD3+ events, relying on the CD3 vs ic-CD137 dot plot, and defining the CD3dim/+ ic-CD137bright population. This approach allowed higher recovery of the events of interest and reliable gating of activated T cells, resulting in 32.8% increase in ic-CD137+ events (calculated by the difference between stimulated and unstimulated, analyzed across four donors, in duplicate, at three time points), with no significant impact on the general cell population numbers (total T cells +0.015%, CD4-CD8-+0.07%, CD4/CD8 ratio -0.01%, not shown). This gating approach was therefore applied throughout the study.

### Determining conditions for resolving activation induced markers and cytokines

CD69 (expressed by CD4+ and CD8+ cells) and CD134 (expressed by CD4+) have been among the first published markers, used in conjunction with CD137, to detect antigen-specific T cells in the context of anti SARS-Cov-2 immune response (Grifoni et al., 2020). We aimed to test the intracellular versus surface staining of these induced markers, with the goal of defining the most reliable approach for measurement of AIM+ T cells when conducted together with intracellular cytokine assesment following antigen-specific activation. As shown in Figure 1A, staining of ic-CD69 strongly correlated with ic-CD137 expression for both CD4+ (row I) and CD8+ cells (row II) and allowed clear resolution of ic-CD137+ elements able to produce cytokines. On the contrary, the use of ic-CD134 to stain CD4+ cells (row III) or s-CD69 to stain CD8+ cells (row IV) displayed a much lower co-expression level with ic-CD137 and failed to capture most of the ic-CD137+ cytokine producing cells. The use of s-CD134 on CD4+ cells resulted in even lower co-expression levels (not shown).

**Figure 1.**
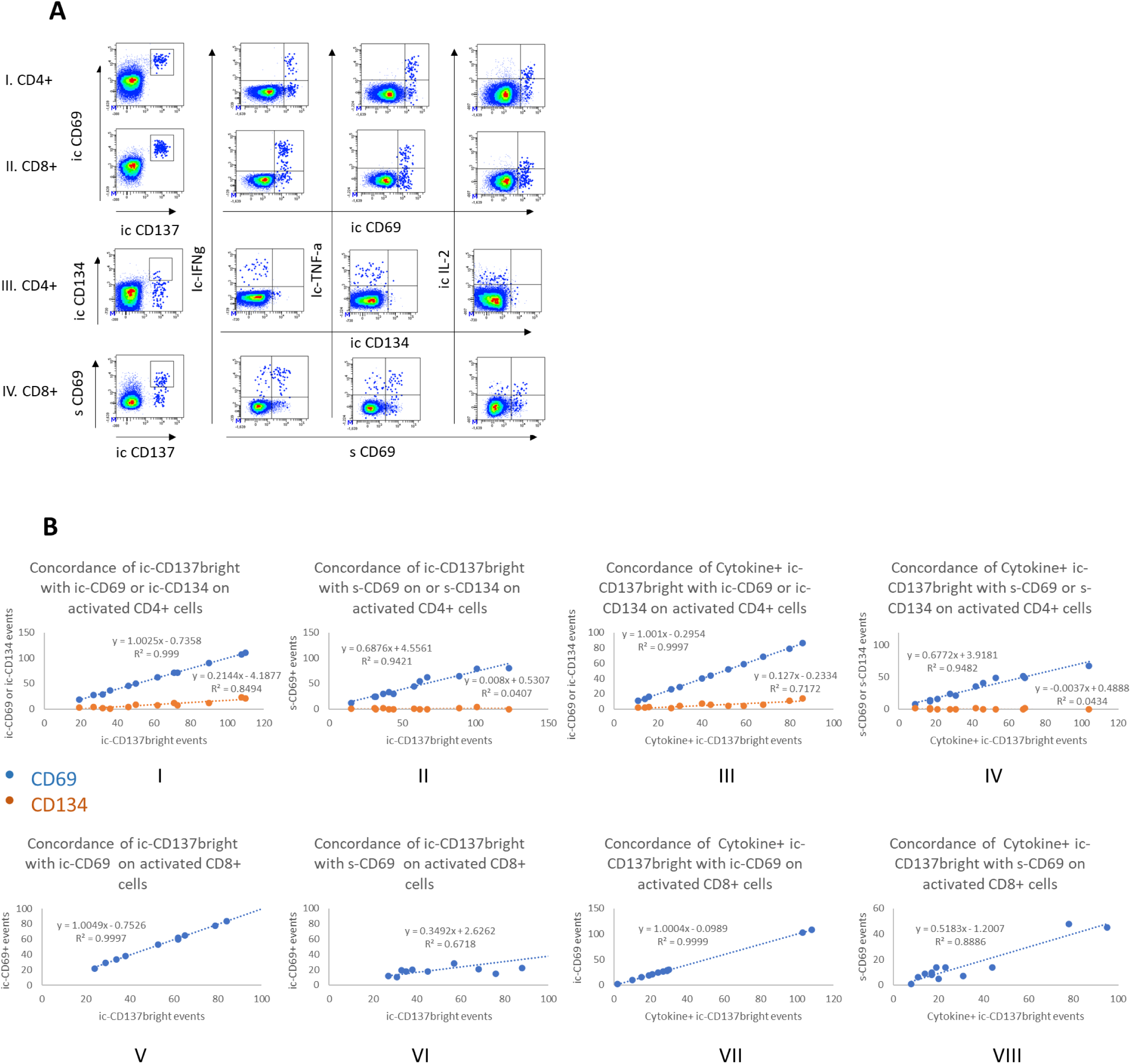
ic-CD69 co-stains with ic-CD137 thus facilitating the measurement of cytokine+ ic-CD137brightCD4+ and CD8+ T lymphocytes A Exemplary plots showing patterns of ic-CD137, s/ic-CD69, ic-CD134, and cytokines on stimulated CD4+ or CD8+ T cells. Rows I and II show ic CD137 CD4+ and CD8+ cells co-stained with ic-CD69: in this approach almost all the CD4+ or CD8+ cytokine producing cells result ic-CD69+. Rows Ill and IV show ic-CD137+ CD4+ and CD8+ cells co-stained with ic-CD134 and s-CD69 respectively: in this approach the majority of CD4+ cytokine producing cells are ic-CD134-and the many of CD8+ cytokine producing cells are s-CD69-. B Analysis of concordance of s/ic-CD69 and s/ic-CD134 staining with total ic-CD137bright CD4+ and CD8+ events and their cytokine producing subset. lc-CD69 is the only marker showing full concordance (Trendline regression slopes =1). Each dot represent a sample: blue dots refer to CD69 staining, orange dot refer to CD134 staining.

Next, we explored the concordance of the three activation induced markers and cytokines for different conditions. Specifically, the co-expression of CD134 or CD69 with ic-CD137 was assessed on four donors at three time points, revealing a clear pattern that is summarized in Figure 1B (graphs I to VIII). ic-CD69 staining perfectly correlated with ic-CD137bright staining on both CD4+ and CD8+ cells. Linear trendlines showed a slope of 1.00 and the correlation coeafficient R^2^ was equal to 0.999 in both cases (Graphs I and V). Conversely, ic-CD134 staining was negative on most of ic-CD137bright CD4+ events having a trendline slope of just 0.21 and R^2^ of 0.85 (Graph I). Interestingly, s-CD69 staining also showed reduced capacity to co-stain with ic-CD137bright on CD4+ and CD8+ cells, with linear trendline slopes of 0.69 and 0.35, respectively, and relative and R^2^ of 0.94 and 0.67 (Graphs II and VI). Notably s-CD134 staining was almost completely negative on ic-CD137bright events on CD4+ subset displaying a trendline slope of 0.04 and R^2^ of 0.04 (Graph II). We then evaluated the capacity of CD69 and CD134 to co-stain with cytokine producing ic-CD137bright CD4+ and CD8+ events. Once again, only ic-CD69 showed a perfect correlation with these events resulting in trendline slopes equal to 1 and R^2^ of 0.999 (Graphs III and VII). s-CD69 on CD4+ or CD8+ correlated to a lower extent with cytokine producing ic-CD137bright elements and s-CD134 did not correlate at all (Graphs IV and VIII).

### Analysis of real AIM+ T cells

We further assessed the effectiveness of s/ic-CD134 and s/ic-CD69 to detect AIM+ T cells in conjunction with ic-CD137, being the percentage of real (r) AIM+ elements calculated as the difference between stimulated and unstimulated samples. As displayed in Figure 2A, ic-CD69 in conjunction with ic-CD137 allowed a higher counting of r-AIM+ CD4+ cells in comparison to ic-CD134. In parallel, ic-CD69 performed better than s-CD69 when either was used in conjunction with ic-CD137 to quantify CD8+ r-AIM+ cells. In the same experiment the effect of different incubation times was explored on the percentages of r-AIM+ T cells (total or cytokine producing fraction) without displaying a clear time-dependent trend. For this reason, we selected the intermediate time of 20 hours of incubation with BFA, and ic-CD69 was indeed designated as the best companion activation marker to be used in conjunction with ic-CD137 for defining r-AIM+ CD4+ or CD8+ T cells.

**Figure 2.**
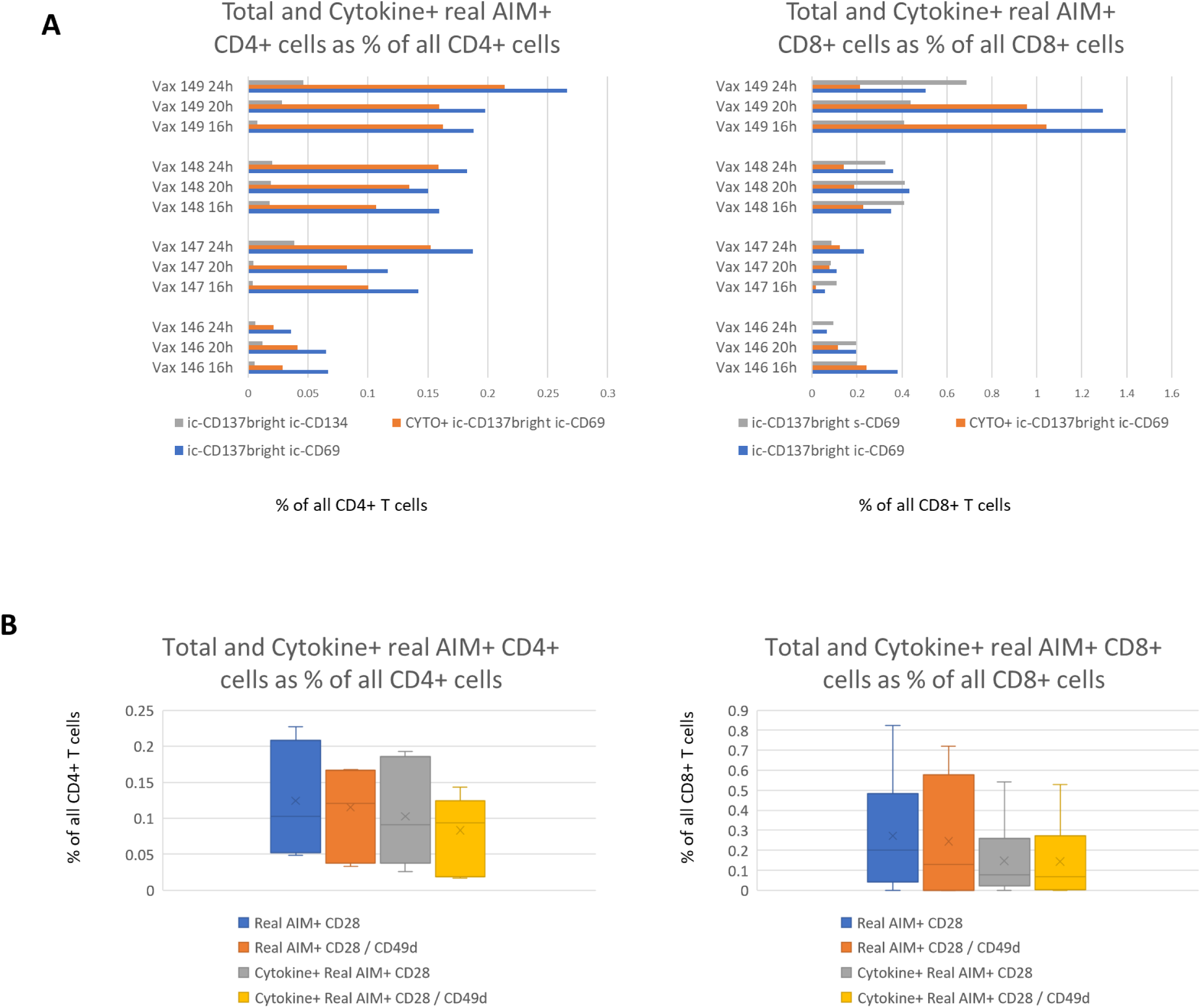
CD4+ and CD8+ real AIM+ assessment based on incubation time and co-stimuli A Graphs represent the percentage of Total and Cytokine+ real AIM+ CD4+ and CD8+ T cells calculated as the difference between stimulated and unstimulated samples. Real AIM+ cells have been evaluated based on different activation phenotypes being for CD4+ cells: ic-CD137bright and ic-CD134+ (gray bars, left graph); ic-CD137bright and ic-CD69+ (blue bars, left graph); Cytokine+ ic-CD137bright and ic-CD69+ (orange bar left graph). With regard to CD8+ cells, activation phenotypes were: ic-CD137bright and s-CD69+ (gray bars, right graph); ic-CD137bright and ic-CD69+ (blue bars, right graph), Cytokine+ ic-CD137bright and ic-CD69+ (orange bars, right graph). The use of ic-CD137 and ic-CD69 results in higher percentage of real AIM+ retrieval for both CD4+ and CD8+. Variation in incubation times in the presence of Brefeldin A from 16 to 24 hours do not lead to clear trends in the percentage of real AIM positive cells. B Graphs represent the percentage of total and cytokine producing real AIM+ out of CD4+ and CDS+ population to compare the costimulatory affect of CD28 only versus CD28 and CD49d. The addition of CD49d does not result in significant changes of real AIM+ events detection, producing or not cytokines. Evaluation was performed on four donors, three of which were assessed in duplicate.

Considering the poor performance of ic-CD134 in comparison to ic-CD69 to detect r-AIM+ CD4+ T cells, we also decided to rule out whether the result was clone dependent. We compared the previously used clone L106 with clone Act35, which is also referenced in the literature as suitable for AIM testing (Grifoni et al., 2020). We activated PBMCs from three healthy donors vaccinated with BNT162b2 and analyzed the percentages of co-expression for ic-CD134 and ic-CD137. As shown in Supplemental Figure 2, ic-CD134 clone Act35 displays higher level of co-expression with ic-CD137 ranging from 14 to 24% versus the lower 9.9 to 19.4% of clone L106. Indeed, despite higher co-expression with ic-CD137, even ic-CD134 Act35 clone shows quite low concordance in comparison with the high concordance previously described for ic-CD69.

As a further fine-tuning step, having the aim to optimize the co-stimulation of T cells, we verified if the use of anti-CD49d in conjunction with anti-CD28 monoclonal antibodies (mAbs) may help to quantify higher percentages of detectable r-AIM+ cells and their cytokine producing subset. We perfomed the comparison on the same four COVID-19 reactive samples previously used in the kinetic experiments, assessing three out of four in duplicate independent experiments. We found that adding CD49d did not result in substantial change in results, and thus it was excluded from further evaluation (Figure 2B).

### Replicate experiments with multiple donors

All the above-mentioned experiments, with the exception of CD134 clone comparison, have been performed on a single batch of previously aliquoted and frozen PBMCs from each of the four vaccinated healthy donors. This allowed cross-experiment comparison in fine tuning the protocol but introduced some restriction on the number of cells used in each experiment (10^6^/each sample). Under these conditions we compared three or four independent tests for each donor (Figure 3A-B), which showed some inter-experiment variability. Sample *Vax146* was a low responder for CD4+ and CD8+ AIM+ cells; *Vax147* was a good responder for AIM+CD4+ and low responder for AIM+CD8+; *Vax148* and *Vax149* were good responders with some variations. We speculate that the observed variability can be attributed to two parallel factors:

**Figure 3.**
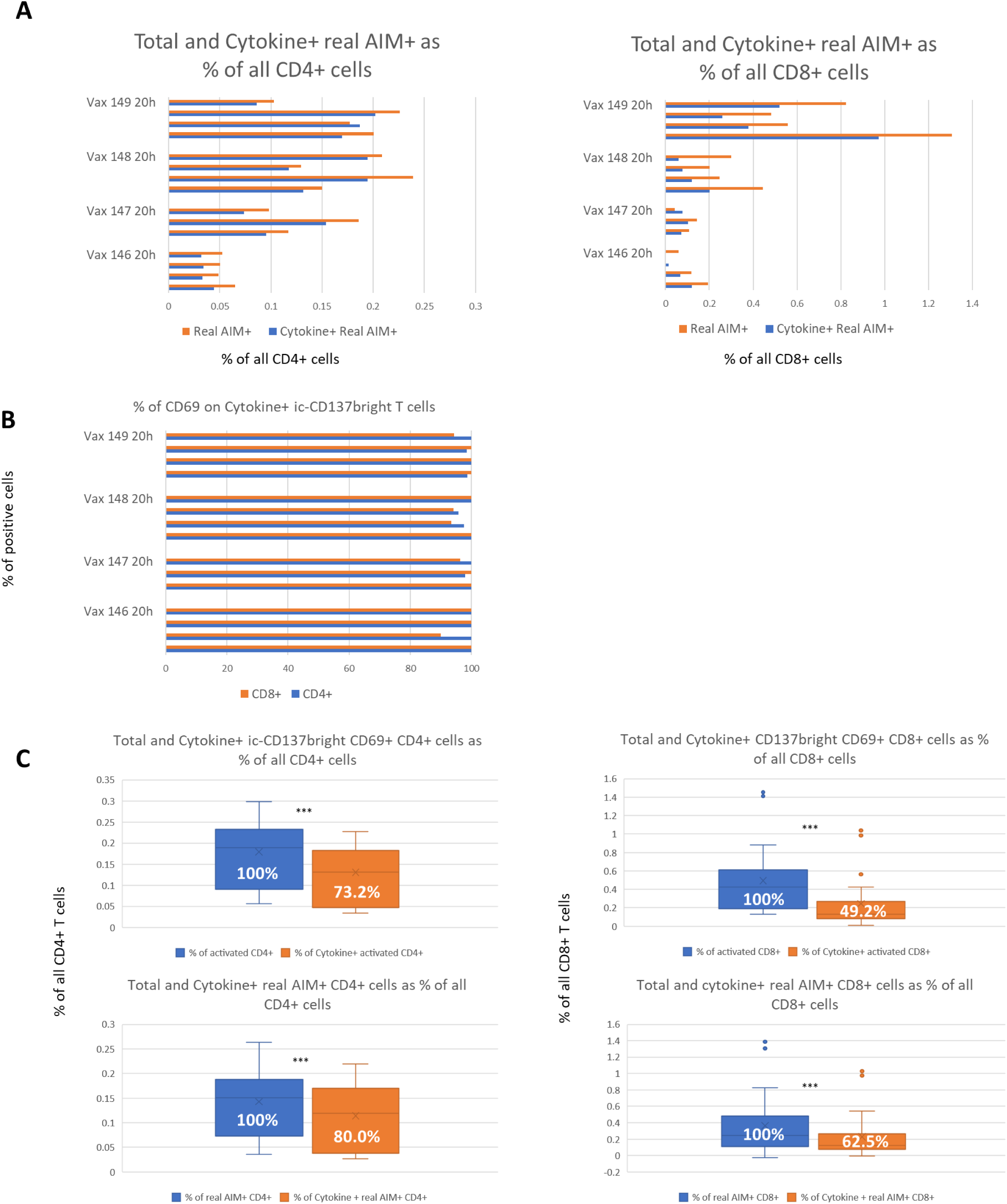
Replicates of AIM assessment and quantification of cytokine producing AIM+ cells A Repeated independent assessment of AIM test in four donors, calculated as the difference in the percentage of AIM+ elements between stimulated and not stimulated samples. The variations observed in different experiments are likely due to the limited number of acquired events and by small differences in background and activation status of each experiment. B Co-expression of ic-CD69 with ic-CD137bright cytokine producing T cells following peptide pool activation. The graph highlights the almost complete overlap between cytokine positivity and expression of ic-CD69. C Comparison between total activated (ic-CD137bright ic-CD69+) and real AIM+ CD4+ and CD8+ with their cytokine producing counterpart. Activated CD4+ or CD8+ T cells are calculated based on stimulated samples only, whereas ‘real AIM+’ T cells and cytokine producing counterpart are calculated as the difference between activated and non activated samples. Y axes refer to percentage of all CD4+ or CD8+ cells, whereas numbers displayed within the graph bars refer to the percentage of cytokine producing fraction of cells with respect of the total activated one.

i. the relatively low number of activated events collected by recording 75,000 live singlet CD3+ cells
ii. the need to estimate r-AIM+ events as the result of a difference between stimulated and not stimulated samples.

Despite the observed variability, the data confirm the excellent capacity of ic-CD69 to stain cytokine producing ic-CD137+ T cells. As shown in Figure 3B, ic-CD69 co-stained from 95.8 to 100% of cytokine+ ic-CD137+ CD4+ T cells and from 93.3 to 100% of cytokine+ ic-CD137+ CD8+ T cells, thus confirming the robustness of this ic-marker to help discriminate TCR-activated T cells when used in conjunction with ic-CD137.

In accordance with previous literature, we confirmed that the amount of cytokine producing activated CD4+ and CD8+ T cells was lower in comparison to total AIM+ events (Figure 3C). Looking at the pool of stimulated samples only, cytokine+ ic-CD137bright ic-CD69+ CD4+ cells were nearly 73.2% of total ic-CD137bright ic-CD69+ CD4+ events (Figure 3C upper left graph), whereas cytokine+ ic-CD137bright ic-CD69+ CD8+ were 49.2% of total ic-CD137bright ic-CD69+ CD8+ (Figure C upper right graph). Notably, when the percentage of real AIM+ elements was correctly calculated as the difference between stimulated and non-stimulated samples, the ratios between cytokine-producing and real total AIM+ CD4+ and CD8+ events was increased to 80% and 62%, respectively (Figure 3C lower left and lower right graphs). Interestingly, the numerical variation introduced by appropriate comparison between stimulated and unstimulated samples are influenced by background events in the non-cytokine producing compartment. In fact, real total AIM+CD4+ and CD8+ are both reduced by nearly 20% and 26% when background from unstimulated samples is subtracted, whereas cytokine producing CD4+ and CD8+ are reduced by nearly 15% and 8%, respectively. This confirms that ic-cytokine based detection of antigen-specific T cell activation is slightly underestimated, yet it is also affected by lower background.

### T cell maturation

We then evaluated the maturation pattern of total and ic-CD137+ ic-CD69+ CD4+ and CD8+ T cells, as shown in Figure 4. As expected, a clear shift in the maturation curve is observed comparing total CD4+ or CD8+ subsets with their matched activated counterpart. ic-CD137+ ic-CD69+ CD4+ were enriched for effector memory and CD8+ cells were enriched for terminally differentiated CD45RA+. Interestingly few naïve cells were also detectable, particularly in the CD4+ population, which may be due to bystander activation. These clearly showed a lower level of CCR7 expression in comparison to total naïve counterparts (Supplemental Figure 3). Lower CCR7 expression was also consistently observed in central memory (CM) activated CD4+ and CD8+ T cells in comparison to total parent populations.

**Figure 4.**
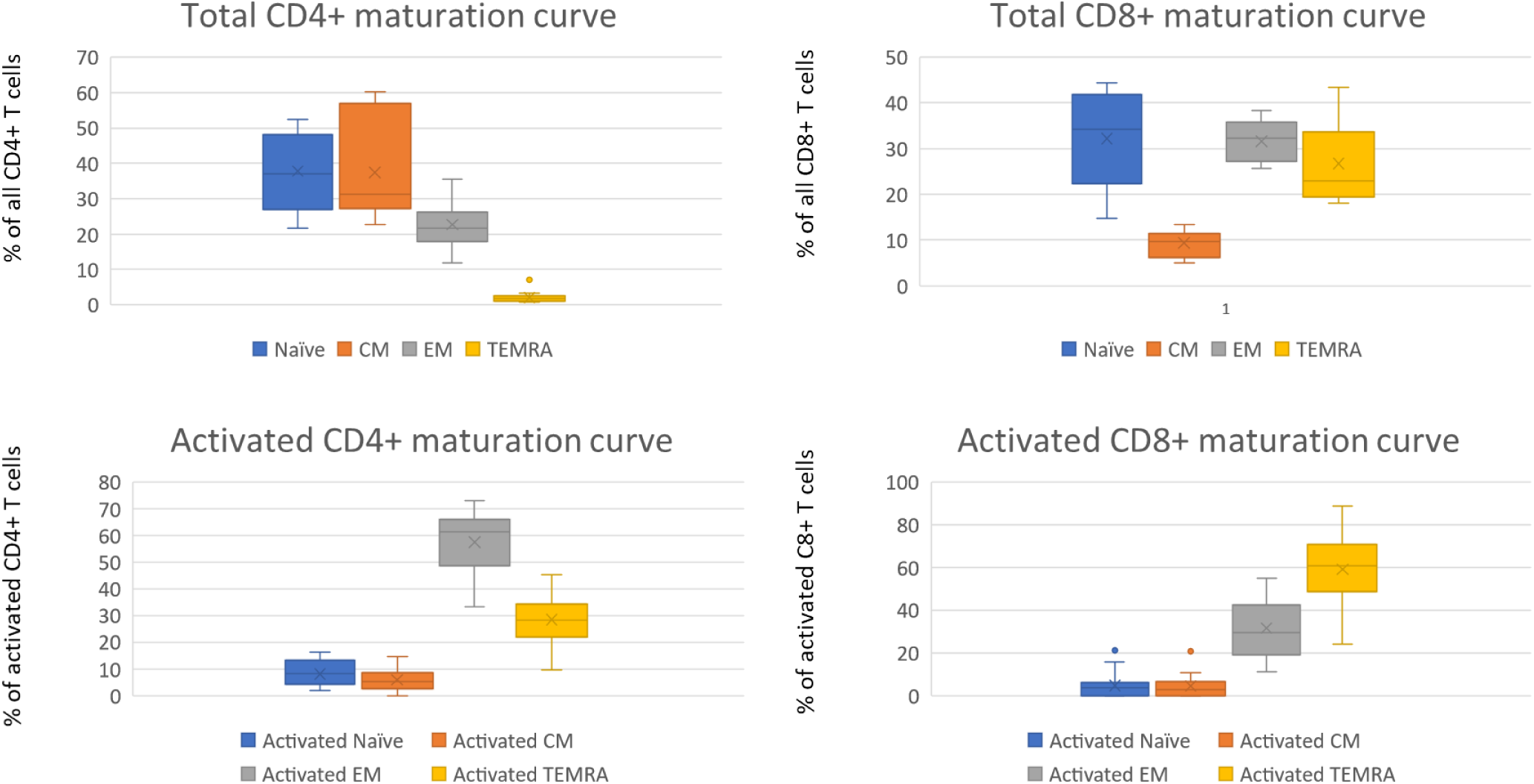
Maturation curve of C04+ and COB+ T cells and their ic-C0137bright ic-C069+ activated counterparts. The most advanced maturation stage is evident for antigen responsive T cells, where activated C04+ T cells are mainly effector memory and activated COB+ T cells are mainly terminally differentiated.

### Final optimization and addition of CXCR5

As a final step we aimed to further optimize the adopted 12-color flow cytometry panel by eliminating the non-informative CD134 marker and including CXCR5 to possibly detect T helper follicular cells, defined as CD4+ CD45RA-CXCR5+ elements (Reiss et al., 2017). To achieve this goal the panel design was slightly reformulated as illustrated in Supplemental Table 2A. The BV421 channel was assigned to CXCR5 for maximum sensitivity, the former CD4 (V450) reagent in this channel was moved in place of CD45RA APC-H7, which was in turn assigned to the BV786 detector, previously dedicated to CD134. These changes left the activation markers and cytokine detection unmodified and allowed great resolution of the T cell maturation curve. The new optimized panel (Supplemental Figure 5) was tested in quadruplicate in parallel with the standard one providing consistent data and highlighting an 8-fold increase of CD45RA stain index calculated on CD4+ cells (not shown). T helper follicular cells were detected in two out of four tested donors, accounting for <10% of r-AIM+ CD4+ cells (not shown).

## DISCUSSION

Multiple methodologies have been described to investigate antigen-specific T cells ex vivo or after in vitro expansion, by direct or indirect interrogation of the T cell repertoire and in combination or not with cell phenotype evaluation. Approaches for ex vivo detection of antigen-specific T cells should rely on minimal in vitro manipulation, so that during short-term stimulation TCR repertoire of antigen-responsive cells is not biased and accurately reflects the clonal dominance hierarchy of the repertoire in vivo.

In the last decade, multiple surface proteins have been established as cytokine-independent surrogates of T cell activation and proven useful to identify and isolate antigen-responsive T cells after TCR engagement by peptide-HLA ligands. These activation markers consist of surface proteins that are rapidly and uniformly upregulated on activated T cells thus allowing the detection of the complete repertoire of antigen-responsive T elements with minimal *in vitro* manipulation, short time of antigen stimulation, and regardless of the functional heterogeneity of reactive T cell themselves.

Based on these premises, the measure of inducible markers on the cell surface in a TCR stimulation-dependent manner, named activation induced marker (AIM) test, is a practical approach to study and monitor antigen-specific T cells. Recently, it has been proposed that AIM testing can be performed in conjunction with intracellular cytokine detection (Braun et al., 2020). In this context, it should be considered that the use of protein secretion inhibitors could negatively impact the expression of activation markers on the cell surface, resulting in an underestimation of the frequency of antigen-responsive T cells. For these reasons, optimizing appropriate conditions for simultaneous AIM testing and cytokine detection is critical.

In the pool of TCR-dependent activation markers to identify antigen-responsive T cells with high specificity, 4-1BB (CD137) is reported to be selectively expressed on all activated CD4+ and CD8+ T cells, and its upregulation is independent of cytokine secretion profile or cell differentiation stage. CD137 expression reaches a maximal level of expression 24 h after *in vitro* stimulation on both CD4+ and CD8+ T cells (Wolfl et al., 2007), (Wehler et al., 2008).

In the design of AIM test assays, preferably different activation markers are used in association to facilitate gating and improve specificity, and several optimized combinations have been reported in the literature to identify preferably antigen-specific CD4 + or CD8 + T cells (Grifoni et al., 2020), (Painter MM. et al, 2021).

Here, we tested ic-CD137 upregulation in association with OX40 (CD134) and CD69. The first is suggested as an inducible marker for CD4+ T cells that has a role in regulating the development of effector and memory stages, while the second is an early activation marker that is frequently linked with cytokine production in intracellular staining (Grifoni et al., 2020). Although it is known that CD69 can be upregulated in a TCR-independent manner (Bremser et al., 2015), it has been also proposed that its use in association with other TCR-dependent activation markers like CD137 may improve sensitivity for the discrimination of TCR-triggered cells (Bacher et al., 2013).

In our hands, intracellular assessment of CD137 and CD69 allowed clear detection of an activated T cell population, composed of both CD4+ and CD8+ T cells, which was evidently increased upon in vitro stimulation with Spike peptide pool. Importantly, both ic-CD137 and ic-CD69 were equally efficient in detecting activated cytokine producing cells (Figure 1 panels A-B).

On the contrary, use of ic-CD134 in conjunction with ic-CD137 was much less effective in both detecting the pool of activated T cells and capturing the cytokine producing T cell fraction (Figure 1). Surface staining of either CD69 or CD134 clearly showed worse results than intracellular staining, as expected due to the protein transport inhibition, which was required to allow intracellular cytokine accumulation. In particular, a much lower amount of cytokine producing ic-CD137+ T cells co-stained with surface CD134 or CD69 thus breaking the key concordance between activation and production of cytokines, which was particularly evident in the case of CD134.

Synergic use of CD137 and CD69 markers for AIM testing is reported in the literature although not in conjunction with intracellular cytokine detection (Geers et al., 2021). In the AIM protocol without evaluation of cytokine production, a better correlation between CD69 and CD134 has been described, which we did not observe here due to different experimental conditions.

Interestingly, we also detected that the fraction of activated cytokine producing CD8+ T cells is characterized by higher ic-CD137 brighteness, thus further emphasizing the good correlation between this marker and T cell activation (Supplemental Figure 4).

Gating of ic-CD137bright CD3dim/+ cells on the relative CD3 vs CD137 plot has been crucial in our analysis approach (Supplemental Figure 1B), allowing the clear and comprehensive gating of activated T cells. The pattern showing selective downregulation of CD3 intensity on ic-CD137bright cells was an aid for unequivocal gate positionng in all the analyzed samples.

The use of CD45RA and CCR7 markers allowed describing the maturation curve of total and activated CD4+ and CD8+ cells. Data clearly indicate a more advanced maturation stage for ic-CD137+ ic-CD69+ T cells that were enriched for effector memory (CD4+) and terminally differentiated (CD8+) subsets. Interestingly, a subset of naïve ic-CD137+ ic-CD69+ CD4+ T cells was also detectable, likely due to bystander activation. Further investigation would be required to properly classify this specific subgroup.

The possibility to detect CD4+ and CD8+ activation with CD137 and CD69 markers, and remove the less informative CD134, allowed addition of CXCR5 to our 12-color flow cytometry panel, thus adding specificity for detecting T follicular helper cells and their antigen-specific subset. Since Tfh cells support antigen-specific B cells in antibody production, this simple addition enables the method to provide insights on the relationship between antigen-specific T cell activation and humoral immunity (Reiss et al., 2017), (Dan, 2017).

The main limitations of the current study are: 1) the experimental model system is rooted in the context of SARS-Cov-2 and 2) the small number of donors and variability. On the first, the study is conducted using vaccinated donors assessed several weeks after BNT62b2 vaccination (Supplemental Table 1) and stimulated with Spike peptides, as well as using control stimulation with PHA, but has not been studied for other T-cell activation conditions. On the second, the study used four vaccinated donors with samples split into several cryopreserved aliquots (10^6^ PBMCs/sample) for comparison across multiple conditions and variability ensued for at least two reasons, i) relatively few activated T cells were detected in each sample, perhaps due to the starting cell numbers and since on average 22.75 weeks had ensued since the last boost (for *Vax 146-149*), such that in most cases another boost would now be recommended, and ii) the introduction of more random noise by needing to subtract background activation from stimulated activation to properly determine the percentage of AIM+ T cells. Recommended improvements for evaluating individual samples in detail are to consider running multiple replicates, multiple timepoints, evaluation closer to time of boost, and increasing the number of processed cells. Nevertheless, taken altogether the data presented here clearly show the optimal performance of intracellular CD137 and CD69 together with intracellular cytokine detection.

In conclusion, we have optimized a robust and sensitive protocol combining AIM testing with intracellular cytokine detection and maturation curve evaluation for Spike-specific T cells. Our approach relies on minimal sample manipulation, relatively low number of cells and an easy-to-perform protocol. This approach is proposed for comprehensive ex vivo evaluation of the T cell mediated immunity. It has been applied for studying responses during SARS-Cov-2 infection or following vaccination, yet it is potentially applicable to any other setting requiring investigation of antigen-specific T cell activation.

## METHODS

Specimen collection. Blood samples have been obtained from BNT162b2 fully vaccinated healthy volunteers within several weeks after the second dose (Supplemental Table 1, for *Vax146-149*, which were most frequently used in this study, the average is 22.75 weeks post-boost) and after informed consent (Supplemental Table 1). Peripheral blood mononuclear cells (PBMCs) were isolated by density gradient cell separation media (Cedarlane) from whole blood collected in heparin tubes (BD Vacutainer^®^, BD) and used after cryopreservation in fetal bovine serum (FBS, Euroclone) supplemented with 10% dimethyl sulfoxide (DMSO, MERCK).

Materials. Antibodies for flow cytometry staining (Supplemental Table 2 B), monoclonal antibodies against CD28 and CD49d costimulatory molecules, BD GolgiPlug™ protein transport inhibitor (Brefeldin A, BFA), BD Stain Buffer supplemented with BSA, and BD Cytofix/Cytoperm™ solution kit were provided by BD.

PepTivator® SARS-CoV-2 S, S1 and S+ for antigen-specific T cell stimulation were provided by Miltenyi Biotec.

Cell culture and activation. Cryopreserved PBMCs were quickly thawed, washed and resuspended at 10^7^/ml in RPMI 1640 complete medium supplemented with 10% FBS and 1x Antibiotic/Antimycotic (Gibco). 10^6^ cells were seeded per well in a 96-u bottom plate and stimulated or not with 1 μg/ml of PepTivator® SARS-CoV-2 Prot_S, Prot_S1, Prot_S+ (Miltenyi Biotech). Monoclonal antibodies anti-CD28 and /or anti-CD49d at 1 μg/ml were added as co-stimulus. After 4h of incubation at 37°C, 5% CO2, the protein transport inhibitor BD GolgiPlug™ (BFA, 1μl/ml) was added in each condition for a further incubation of 16, 20 or 24 hours. Positive controls were stimulated with Phytohemagglutinin (PHA, Thermofisher) at 10μg/ml.

Cell staining. After incubation with BD GolgiPlug™, cells were washed with 100 ul of Stain Buffer to remove culture medium. Staining was performed in 96-well plates. Cells were blocked using BD Fc Block™ Receptor for 10 min at room temperature (RT) and afterward stained 15 min RT with the surface antibody cocktail and viability dye (Table 1). For subsequent intracellular protocol, samples were washed using Stain Buffer and fixed with 200 μl of Cytofix/Cytoperm for 20 minutes at RT. After washing with 200 μl BD Perm/Wash™ buffer, cells were incubated with intracellular antibodies for 20 min at RT (Table 1). Samples were finally washed with Stain Buffer and re-suspended in 400 μl of PBS in 5 ml polystyrene tubes for acquisition with BD FACSCelesta™ flow cytometer equipped with Blue, Violet and Red lasers.

Flow cytometry and data analysis. Cells were acquired on a BD FACSCelesta™ flow cytometer and analyzed using BD FACSDiva™ software (v9.0). Compensation was calculated using single-stained anti-mouse Ig,κ Comp Beads (BD Biosciences). Doublets were excluded by plotting forward scatter area versus forward scatter height. Viable lymphocytes were defined as fixable viability stain neg-low cells. Combinations of cells expressing AIM, maturation markers, and cytokines were determined using appropriate gating strategy (Supplemental Figure 5)

Calculations. AIM+ cells are calculated by subtracting the percentage of CD137+CD69+ events in unstimulated samples from stimulated samples. Parametric data were analyzed by paired Student’s t test (one tail). P values are summarized as follow: * = p<0.05; ** = p<0.005; ***=p<0.0005.

## Data Availability

All data produced in the present study are available upon reasonable request to the authors

---

**SUPPLEMENTAL FIGURE 1.**
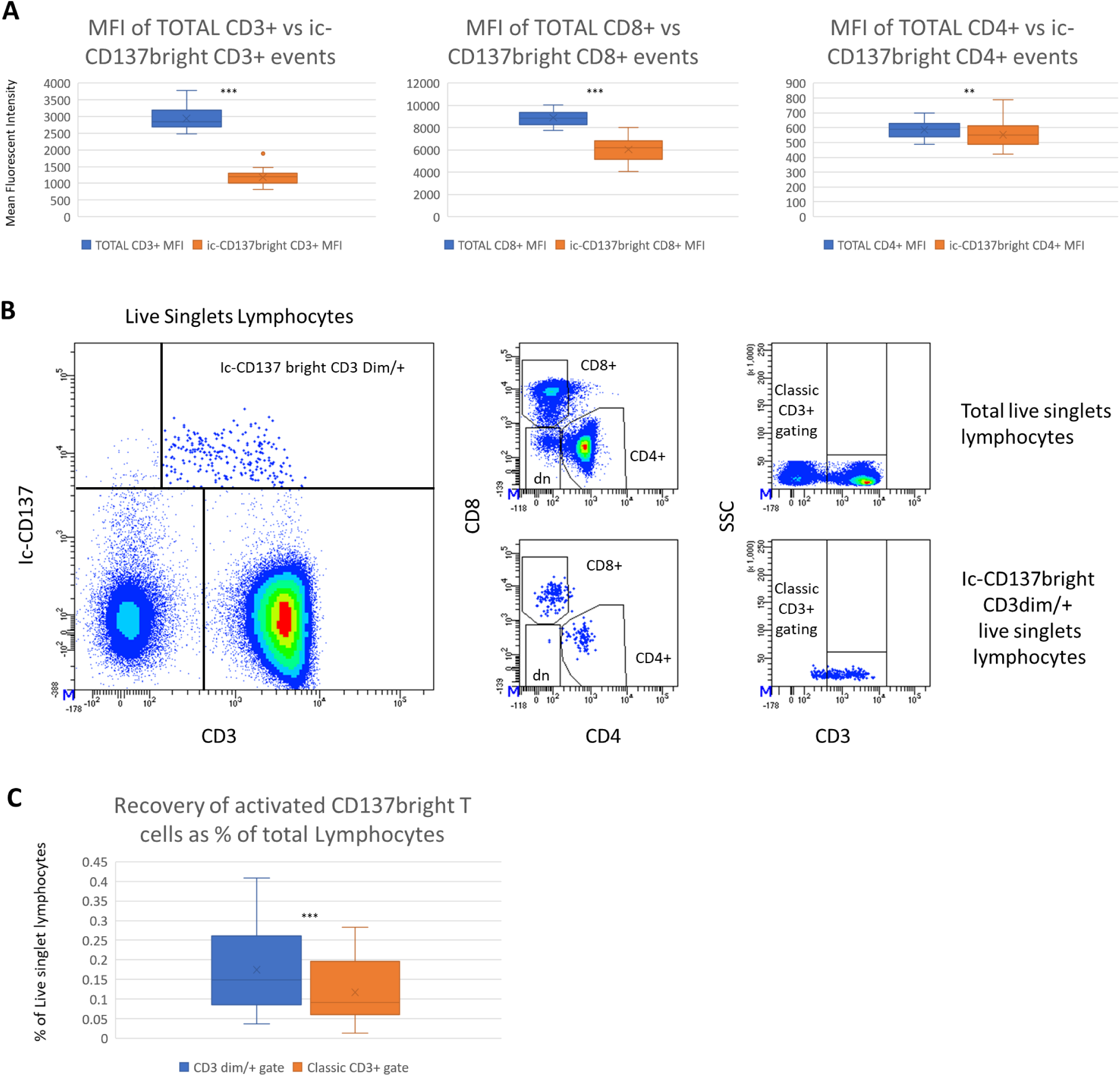
Developing the gating strategy A Decrease of CD3, CD4, CDS Mean Fluorescence Intensity of ic-CD137brightT cells in respect of total CD3+ T cells. B Optimized gating strategy for the ic-CD137bright CD3dim/+ subpopulation. Events of interest are highlighted with large dots. Total CD3+ population results form adding events of upper right and lower right quadrants. Smaller plots display positioning of ic-CD137brightCD3dim/+ events in classical CD4, CD8 and CD3 gates. C Increased recovery of activated T cells (calculated as the difference between stimulated and unstimulated samples) obtained by optimized CD3 gate including ic-CD137bright CD3dim/+ events. The average extent of recovery was equal to 32.8%.

**SUPPLEMENTAL FIGURE 2.**
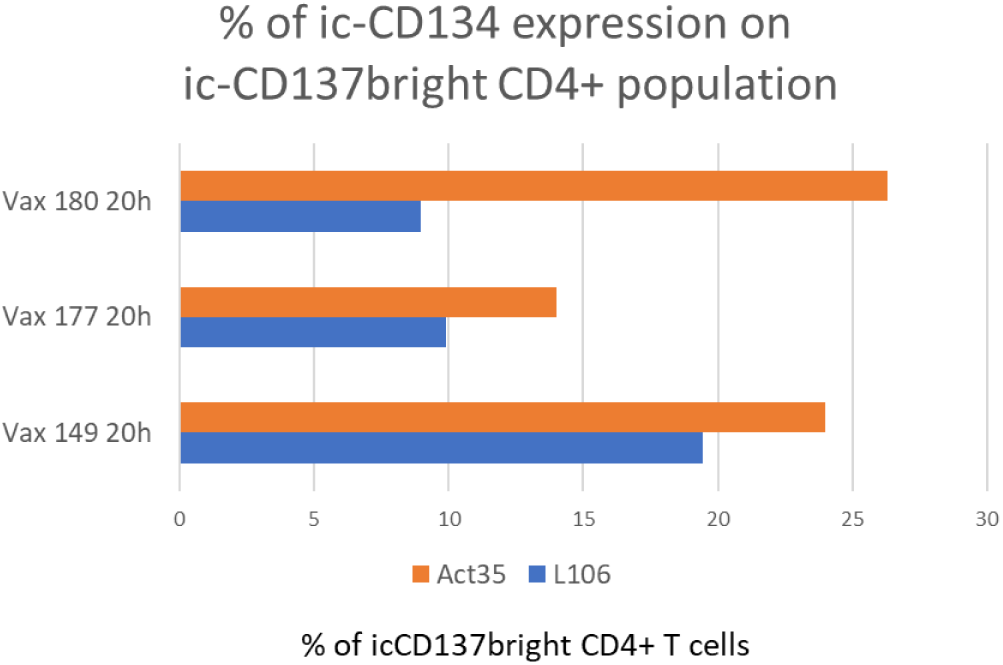
Comparison of ic-CD134 staining level on ic-CD137brightCD4+ population using L106 and Act-35 clones. Three donors have been assessed showing high co-expression percentages when clone Act-35 was adopted.

**SUPPLEMENTAL FIGURE 3.**
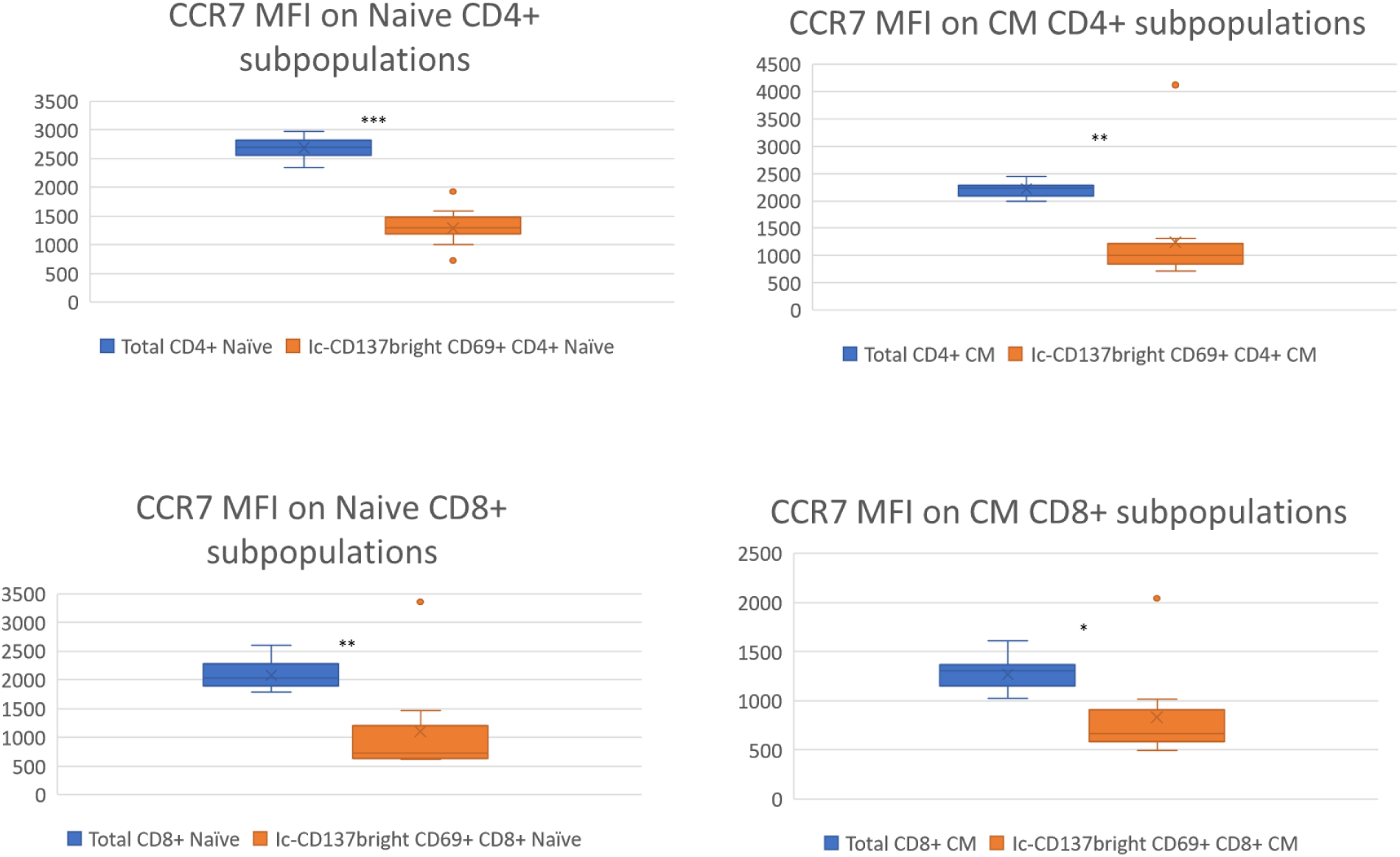
Comparison of CCR7 MFI between total Na’ive and Central memory T cells and their activated CD137+ ic-CD69+ counterpart shows a strong CCR7 downregulation in antigen responding T cells.

**SUPPLEMENTAL FIGURE 4.**
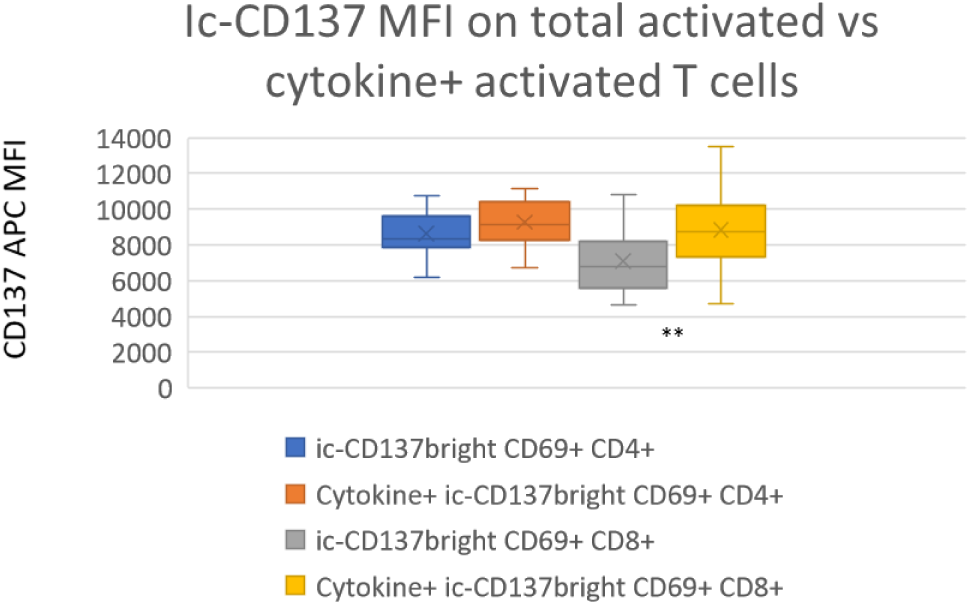
ic-CD137 Mean Fluorescent Intensity was measured for the entire 137bright CD4+ or CDS+ population and compared with the relative cytokine+ CD137bright subpopulation. Cytokine producing ic-CD137bright cells are characterized by higher expression of the marker.

**SUPPLEMENTAL FIGURE 5.**
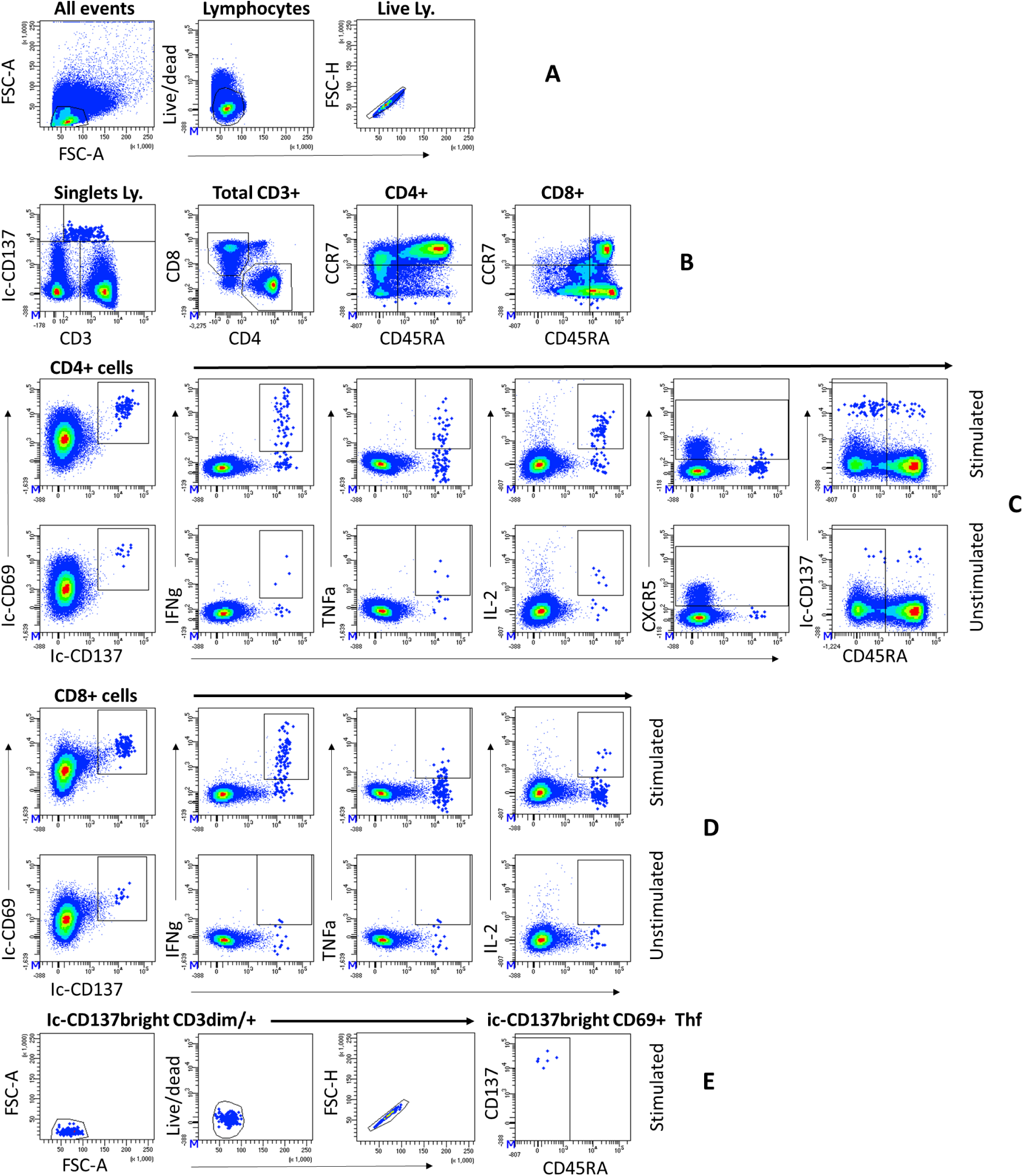
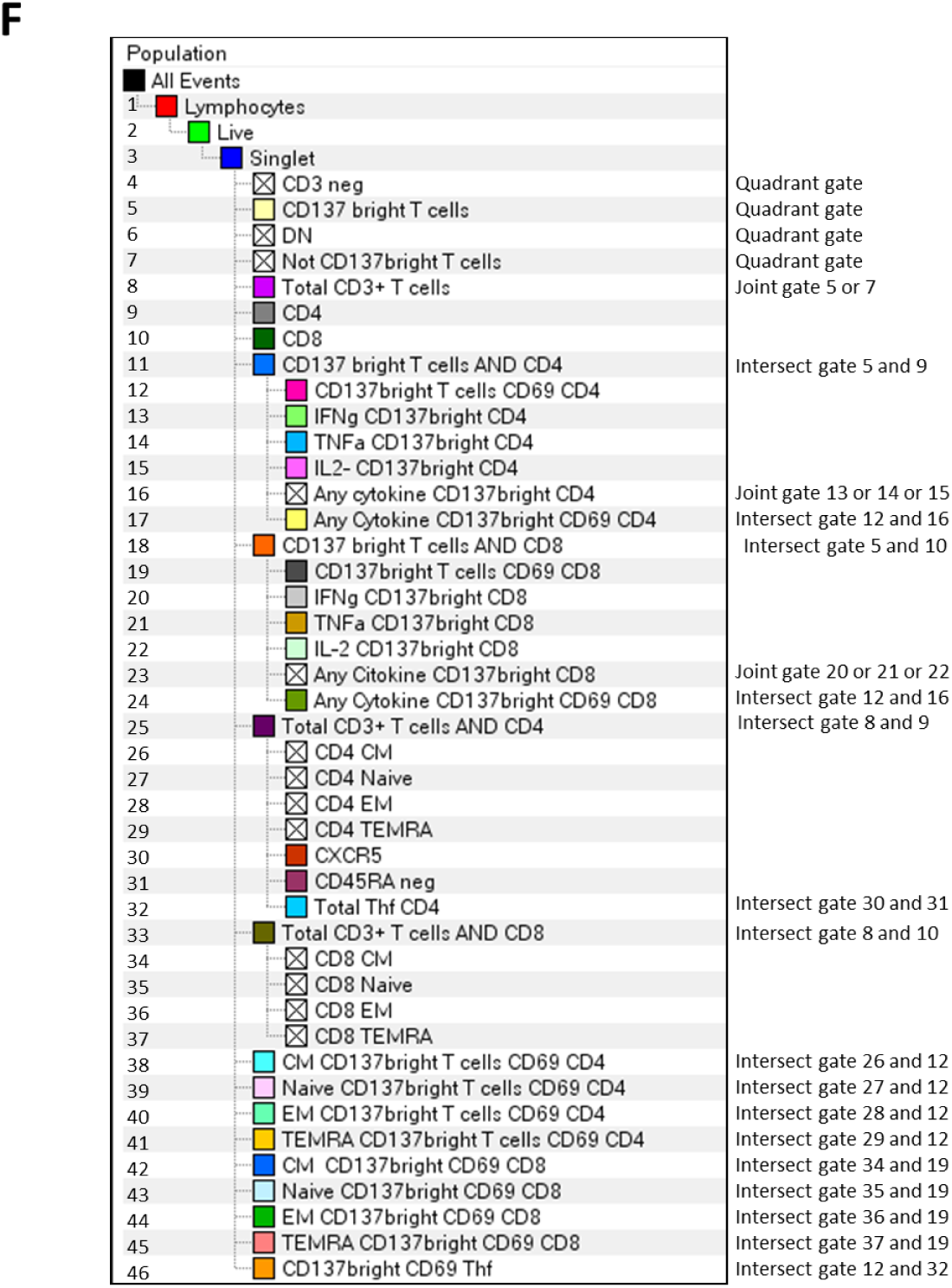
Representative dot plots and relative gating strategy A Live singlet lymphocyte gating B Total CD3 gating by means of the CD3 vs CD137 plot. CD4+ and CD8+ gating and relative maturation curves C Stimulated and unstimulated CD4+ events, including maturation curve and T helper follicular cells D Stimulated and unstimulated CD8+ events, including maturation curve E Verification dot plot to check the correctness of fundamental gate positioning for detecting the ic-CD137bright CD3dim/+ population and the Thf subset

**SUPPLEMENTAL TABLE 1.** DONORS DEMOGRAPHICS.

|  | Sex | Age range | Weeks after 2nd dose of BTN162n2 | Post vaccine COVID pathology | Weeks from last negative swab |
| --- | --- | --- | --- | --- | --- |
| Vax 146 | F | 30-35 | 23 | No | NA |
| Vax 147 | M | 30-35 | 12 | No | NA |
| Vax 148 | F | 60-65 | 24 | No | NA |
| Vax 149 | M | 55-60 | 32 | Yes | 8 |
| Vax 177 | F | 50-55 | 30 | No | NA |
| Vax 180 | F | 40-45 | 38 | No | NA |

**SUPPLEMENTAL TABLE 2.**
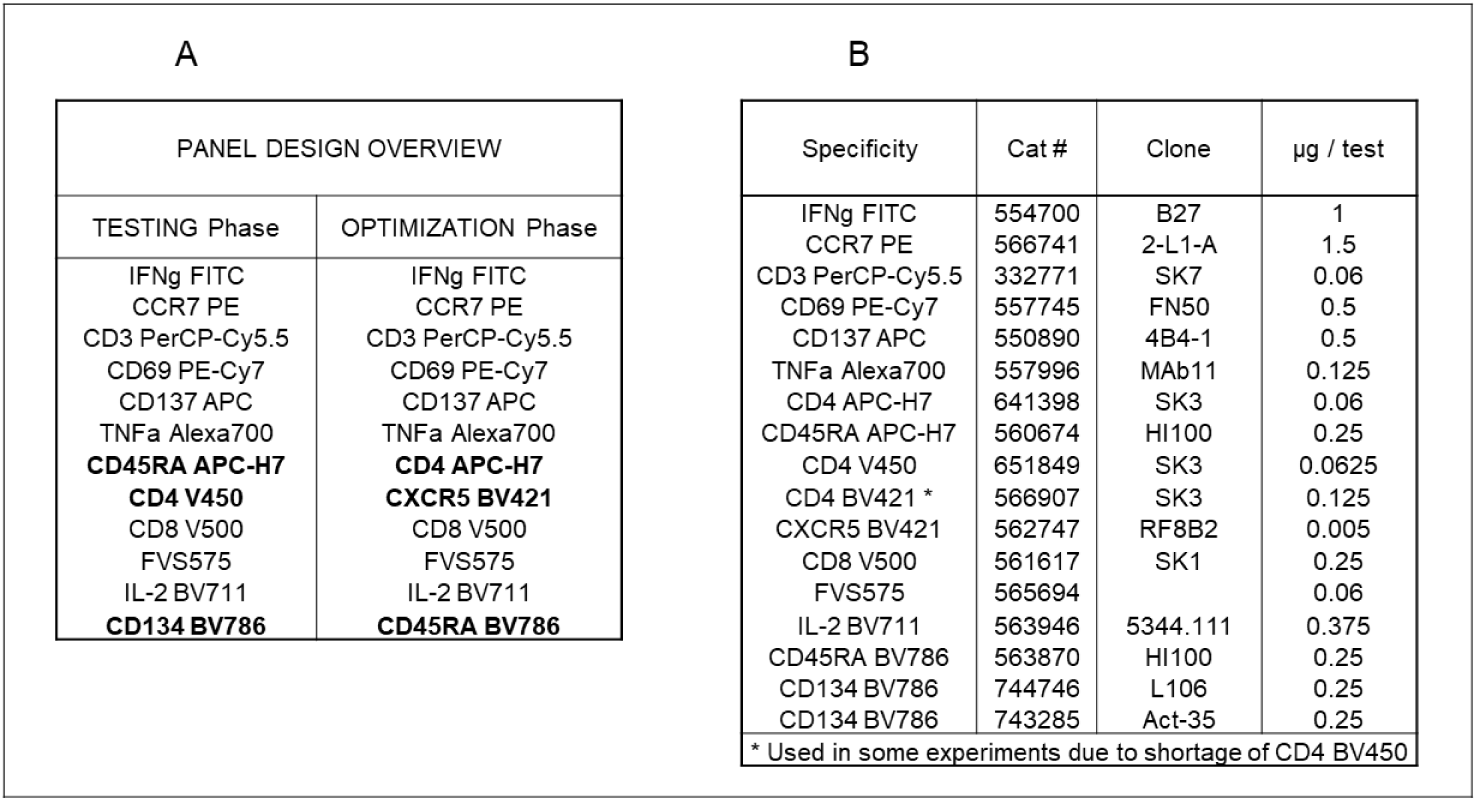
DONORS DEMOGRAPHICS. A) FLOW CYTOMETRYPANEL DESIGN B) REAGENT DESCRIPTION

